# Comprative evaluation of Advanced Oxidation Processes (AOPs) for reducing SARS-CoV-2 viral load from campus sewage water

**DOI:** 10.1101/2022.11.16.22282387

**Authors:** Rinka Pramanik, Narendra Bodawar, Aashay Brahme, Sanjay Kamble, Mahesh Dharne

## Abstract

Although the presence of SARS-CoV-2 fragments in raw sewage water are not much concerning, since it is a new pathogen and its fate in the environment is poorly understood; therefore efforts are needed for their effective removal. In under-developed countries with poor sewersheds and sanitation practices, the raw sewage water might come in contact with rivers and other water bodies and is generally used by the population for various purposes including drinking water. Hence it is important to properly treat sewage water to reduce public health risks, if any. Our study evaluated various advanced oxidation processes (AOPs) for disinfection of SARS-CoV-2 from sewage water collected from the academic institutional residential campus. The present study is the first report showing hydrodynamic cavitation (HC) used to reduce the SARS-CoV-2 viral load from sewage water. Additionally, we have also evaluated hybrid techniques like HC/O_3_, HC/O_3_/H_2_O_2_, HC/H_2_O_2,_ O_3_/UV, UV/H_2_O_2_, UV/H_2_O_2_/O_3,_ and O_3_/H_2_O_2_ for the minimization of the SARS-CoV-2 viral load from sewage water. The sewage water treatment techniques were evaluated based on its viral concentration-reducing efficiency by comparing it with the same raw sewage water sample. However, ozone alone and its combination with other disinfecting techniques (like HC, UV, and H_2_O_2_) showed >95% SARS-CoV-2 specific RNA-reducing efficiency (also known as viral load). The AOPs treated sewage water was subjected to total nucleic acid isolation followed by RT-qPCR for viral load estimation. Interestingly, all sewage water treatment techniques used in this study significantly reduces both the SARS-CoV-2 viral load as well as PMMoV (faecal indicator) load.

## 1. Introduction

The first detection of the SARS-CoV-2 virus responsible for causing COVID-19 infection was found in Wuhan, China, in December 2019 (Jalali Milani and Nabi Bidhendi, 2022; Kitajima et al., 2020; Serra-Compte et al., 2021). The COVID-19 outbreak was declared a public health emergency of international concern by the World Health Organization (WHO) on 30^th^ January 2020. The SARS-CoV-2 virus is mainly transmitted through tiny respiratory droplets (Nasir et al., 2021). COVID-19-infected patients, both symptomatic as well as asymptomatic excrete the SARS-CoV-2 virus through faeces and other body secretion (sputum, saliva, urine) that are released via the restroom or lavatory and is introduced into the sewage treatment plant (STP) (Foladori et al., 2022; Gwenzi, 2021; Jalali Milani and Nabi Bidhendi, 2022; Serra-Compte et al., 2021) SARS-CoV-2 RNA can survive in wastewater for around eight days, and it can remain inactive at 4° C for approximately 19 days (Beattie et al., 2022; Wang et al., 2005). Therefore, sewage water from the inlet of STPs can be used to extract SARS-CoV-2 viral RNA, and its presence and quantification can be determined using RT-qPCR. Wastewater-based epidemiology (WBE) estimates the prevalence of diseases and works as an early warning system for population-wide infectious diseases (Hellmér et al., 2014; Saguti et al., 2021; Sims and Kasprzyk-Hordern, 2020). It was already proven to be an effective tool during the Influenza A (H1N1) outbreak and other studies like the presence of Norovirus, Hepatitis A and E virus, adenovirus, and rotavirus (Hellmér et al., 2014).

The water released from the outlet of STPs plays an essential role in protecting public health as it has been used for irrigation, recreational purposes, or discharged in rivers (Foladori et al., 2022; Kokkinos et al., 2020). The presence of SARS-CoV-2 genetic material influences the quality of water in many ways. Such sewage water can cause many public health risks and environmental issues if released without proper treatment from STP’s outlet to the environment. Hence, an effective treatment method for reducing viral load from sewage water is crucial for the environment (Foladori et al., 2020; Kokkinos et al., 2020; Serra-Compte et al., 2021). Many studies demonstrate the presence of SARS-CoV-2 throughout the STPs as well as in treated wastewater around the globe like India, China, Paris, Germany, Japan, USA, Italy, and Spain (Betancourt et al., 2021; Giacobbo et al., 2021; Hellmér et al., 2014; Miyani et al., 2020; Serra-Compte et al., 2021; Sinclair et al., 2008; Spurbeck et al., 2021; Wang et al., 2005; Weidhaas et al., 2021). Primary or secondary STP treatments cannot wholly remove SARS-CoV-2 RNA from raw sewage water; hence tertiary or advanced treatment techniques must be investigated (Serra-Compte et al., 2021). As the primary treatment involves the removal of suspended solids, the complete removal of SARS-CoV-2 RNA is unachievable in this stage (Abu Ali et al., 2021). Besides, the virus can easily survive the primary disinfection techniques because of their size; therefore, chemical disinfection is the alternate approach for eliminating viruses (Nasir et al., 2021). Mainly chlorine-based disinfection strategies are used to remove SARS-CoV-2 RNA from sewage water (Zhang et al., 2020). However, the generation of eco-toxic by-products, such as chloroform, halo acetic acids, and trihalomethanes, sets a significant limitation to this method. Other disinfecting techniques have their merits and demerits, as they are not equally effective for inactivating viral particles (Al-Hazmi et al., 2022; Gholipour et al., 2022; Rex and Chakraborty, 2022) Therefore, it is crucial to explore and develop a proper treatment strategy for contaminated sewage water in battling this kind of pandemic situation.

Advanced oxidation processes (AOPs) are the latest wastewater treatment techniques that efficiently disinfect viruses by generating oxidant species (Kokkinos et al., 2021). The fundamental mechanism for the degradation/disinfection of viruses by AOPs occurs via the generation of Hydroxyl (OH^•^) radicals. Hydroxyl (OH^•^) radical has an oxidising potential of 2.8V (pH – 0) and 1.95V (pH 14) vs SCE (saturated calomel electrode, the most used reference electrode) (Tchobanoglous et al., n.d.). The OH^•^ has a nonselective behaviour and reacts rapidly with a wide range of species, with rate constants in the 10^8^-10^10^ M^-1^ s^-1^ range. Organic pollutants/ viruses get attacked by hydroxyl radicals via four fundamental mechanisms: radical combination, electron transfer, hydrogen abstraction and radical addition (Deng and Zhao, 2015). When hydroxyl radical reacts with organic compounds, they produce carbon-centred radicals (R or R-OH), which can be further converted to organic peroxyl radicals (ROO) using O_2_. All the radicals continue to react, forming more reactive species like H_2_O_2_ and superoxide (O_2_^•^), which leads to the complete mineralisation of pollutants or disinfection of viruses from water bodies. The possible reaction mechanisms of each disinfecting technique have been summarised in Table 1.

**Table 1:**
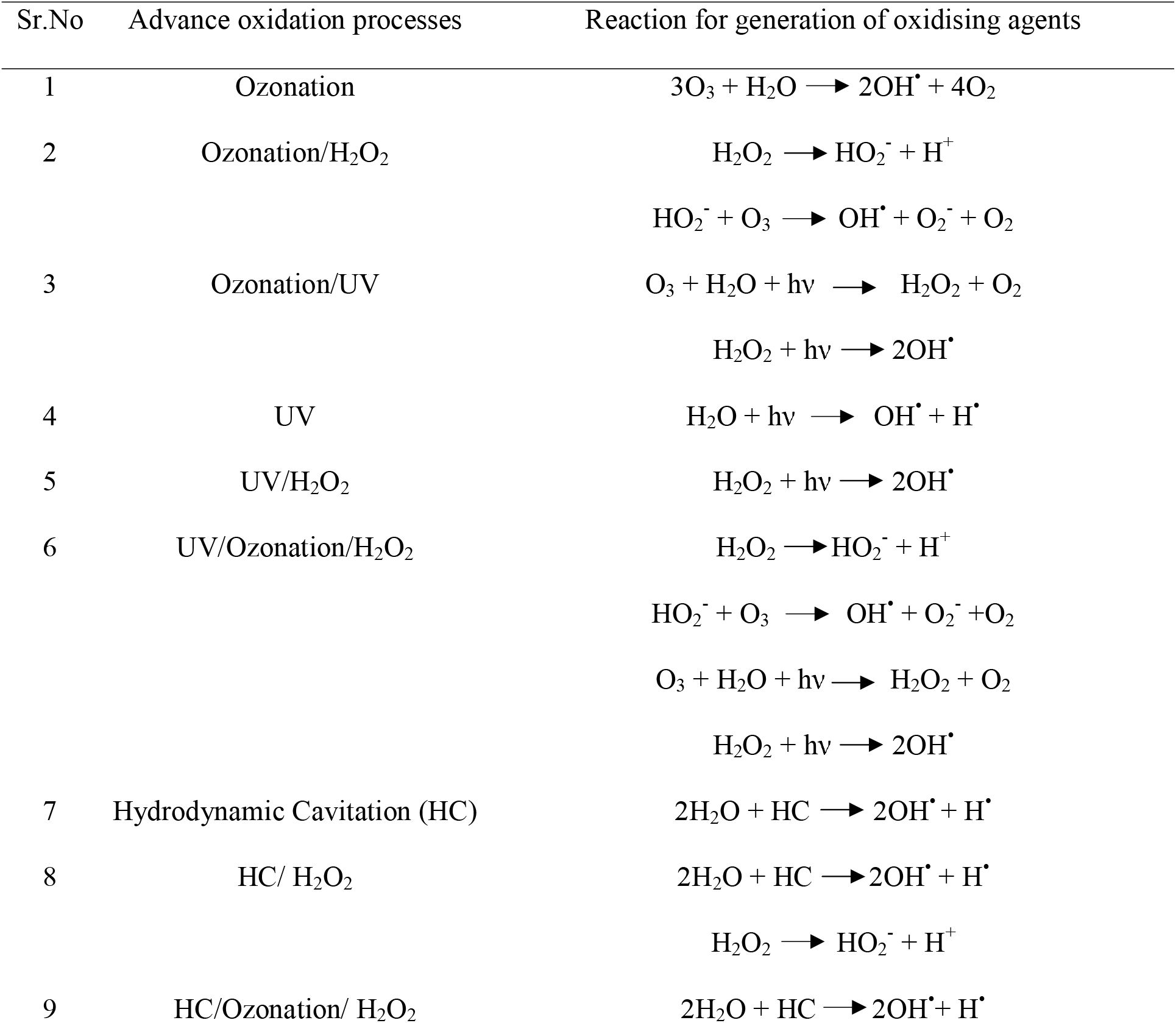
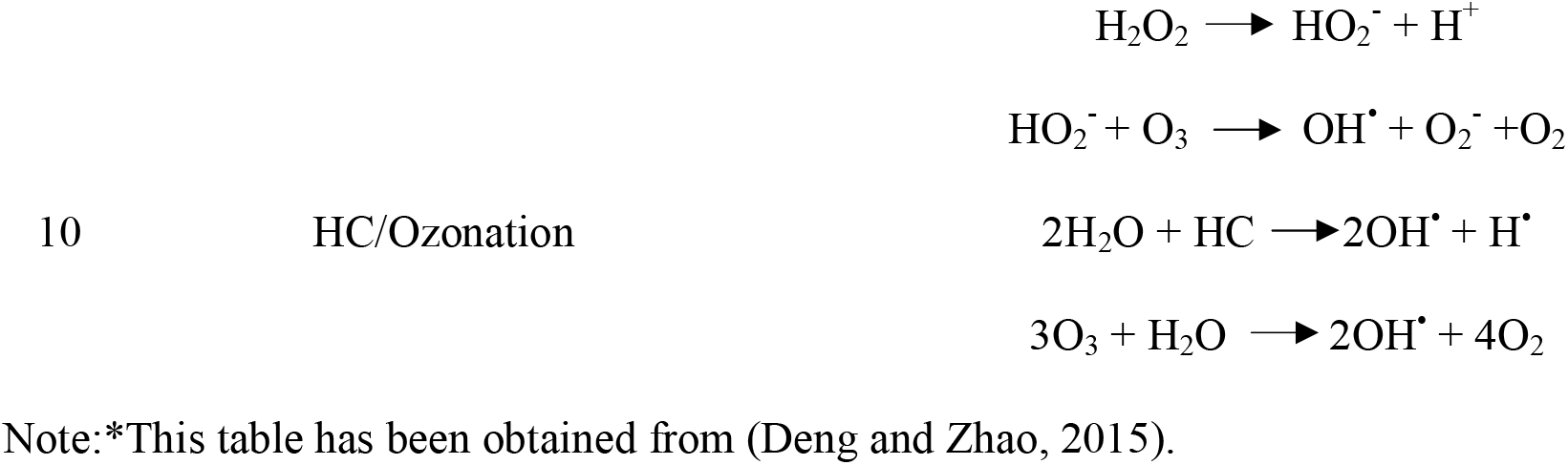
Different AOPs and their possible reaction mechanism for disinfection of SARS-CoV-2*.

This study aims to examine and evaluate advanced oxidation processes (AOPs) and their hybrid combinations for disinfecting SARS-CoV-2 from sewage water. The AOPs used in this study are ozonation, ultraviolet radiation (80 W lamp of wavelength 254 nm), hydrodynamic cavitation (HC), as well as their hybrid techniques like HC/O_3_, HC/O_3_/H_2_O_2_, HC/H_2_O_2,_ O_3_/UV, UV/H_2_O_2_, UV/H_2_O_2_/O_3_ and O_3_/H_2_O_2_ for reducing the SARS-CoV-2 viral load from sewage water. The raw sewage water sample and AOPs treated sewage water samples were subjected to RT-qPCR to discern SARS-CoV-2 viral load. Subsequent comparison of the viral load of treated sewage water with that of raw sewage water, giving six best treatment techniques were shortlisted for further experimentation. Additionally, the effect of ozone dose on these above techniques was studied to establish the optimum ozone dose required. Further selected six best techniques were used to treat three distinct sewage water samples collected on different dates to validate their efficacy for disinfection of SARS-CoV-2 (Fig 1).

**Fig. 1.**
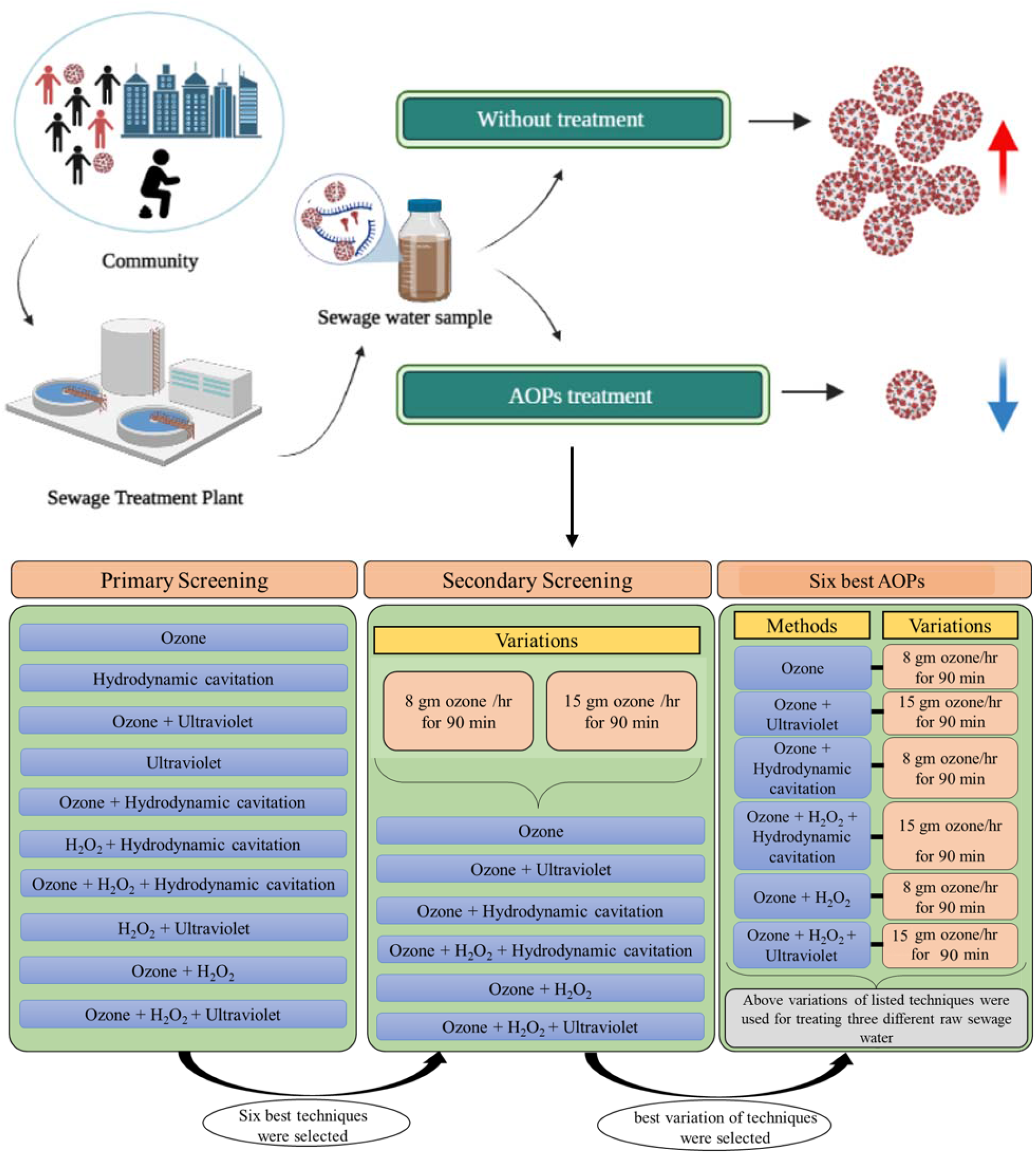
Experimental scheme illustrating treatment of raw sewage water samples (from STP) with different treatment techniques for reduced SARS-Cov-2 viral load. (Image: BioRender.com).

The sewage water treatment techniques would be most effective if they also reduce faecal matter and other enteric viruses from outlets of STPs. Therefore, during the final screening of raw sewage water treatment techniques, we also investigated the reduction of Pepper mild mottle virus (PMMoV). It is the most prevalent virus (up to 10^9^ viruses/gram dry stool) in human faecal samples, as it is a plant virus with the dietary origin of humans from various pepper species (Rosario et al., 2009; Symonds et al., 2018; Zhang et al., 2006). Therefore, it is counted as a viral indicator of faecal contamination in sewage water because of its relatively stable nature and ability to move through the gut quickly (Rosario et al., 2009).

## 2. Material and Methods

### 2.1 Experimental Setup

#### 2.1.1 Setup for Hydrodynamic cavitation and hybrid hydrodynamic cavitation technique

The experimental setup used for hydrodynamic cavitation and hybrid hydrodynamic cavitation for disinfection of SARS-CoV-2 from sewage water is shown in Fig. 2. The cavitation setup consists of an effluent holding tank (EHT) having a capacity of 5 litres, a centrifugal pump (P) power rating of 2 kW (CNP YE2-80M1-2: 2830 rpm, 415V, 50 Hz A.C.), ½ inch control valves (ball valves) and pressure gauges PG1 and PG2. The venturi throat diameter (6mm) and orifice (3 mm) are used as cavitation devices in the present setup. The sewage water was circulated from the EHT through the line consisting of a venturi or orifice device and back into the EHT using a positive displacement pump, and a bypass line was provided to control the circulation flow rate if required. The temperature of sewage water in the EHT tank is controlled by circulating the cold water through helical coils submerged in the EHT tank. The entire setup was fabricated using stainless steel 316. This experimental setup was used to disinfect SARS-CoV-2 from the sewage water using hydrodynamic cavitation and hybrid hydrodynamic cavitation techniques such as HC/O_3_, HC/O_3_/H_2_O_2_, and HC/H_2_O_2._

**Fig. 2.**
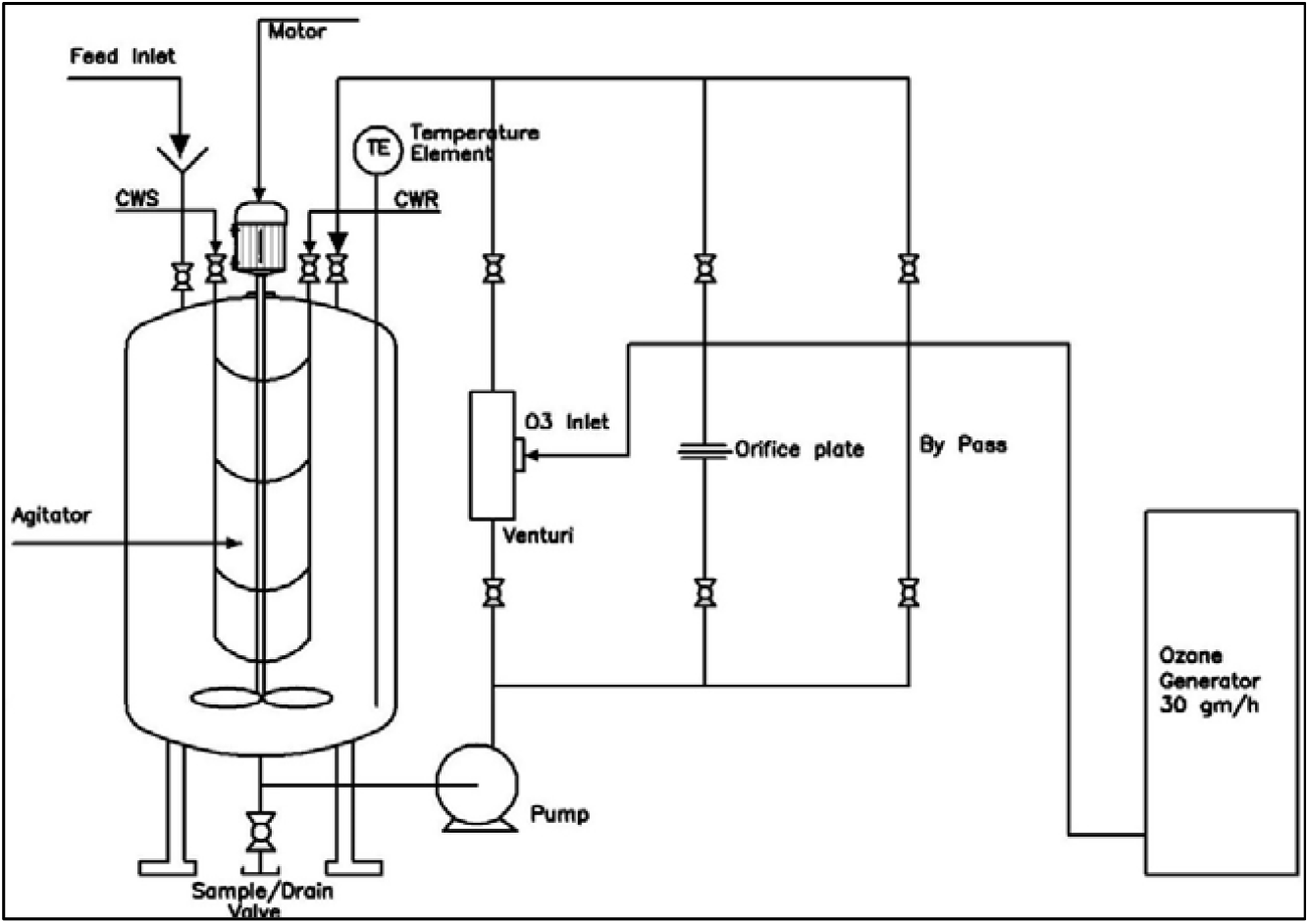
Setup for hydrodynamic cavitation and its hybrid techniques.

#### 2.1.2 Setup for Hybrid Photocatalytic and Ozonation processes

As shown in Fig. 3, an annular glass reactor comprising a quartz candle placed inside the annular glass reactor was used for disinfection of SARS-CoV-2 from raw sewage water. The volume of the glass reactor is 1 litre, with an annular effective volume of 500 ml. The disinfection of SARS-CoV-2 takes place in the annular volume formed between the glass rector and the quartz candle placed in it. The required ozone flow is introduced from the bottom of the glass reactor through a ring air sparger. Ozone generator supplied by Ozonics India Ltd, Pune, India, was used in the present experimental work. It can be operated upto a maximum current of 3 A. The generator produces ozone using dry air as a feed gas. The ozone generator was operated within a current range of 0.08 A to 0.15 A, producing ozone at a flow rate of 8 gm/hr and 15 gm/hr, respectively. The quartz candle is equipped with a cooling water inlet and outlet to maintain the temperature of the sewage water in the reactor. The hollow inner space inside the quartz candle is used to place the UV lamp. The UV lamp having 80 W capacity and wavelength of 254 nm was used.

**Fig. 3.**
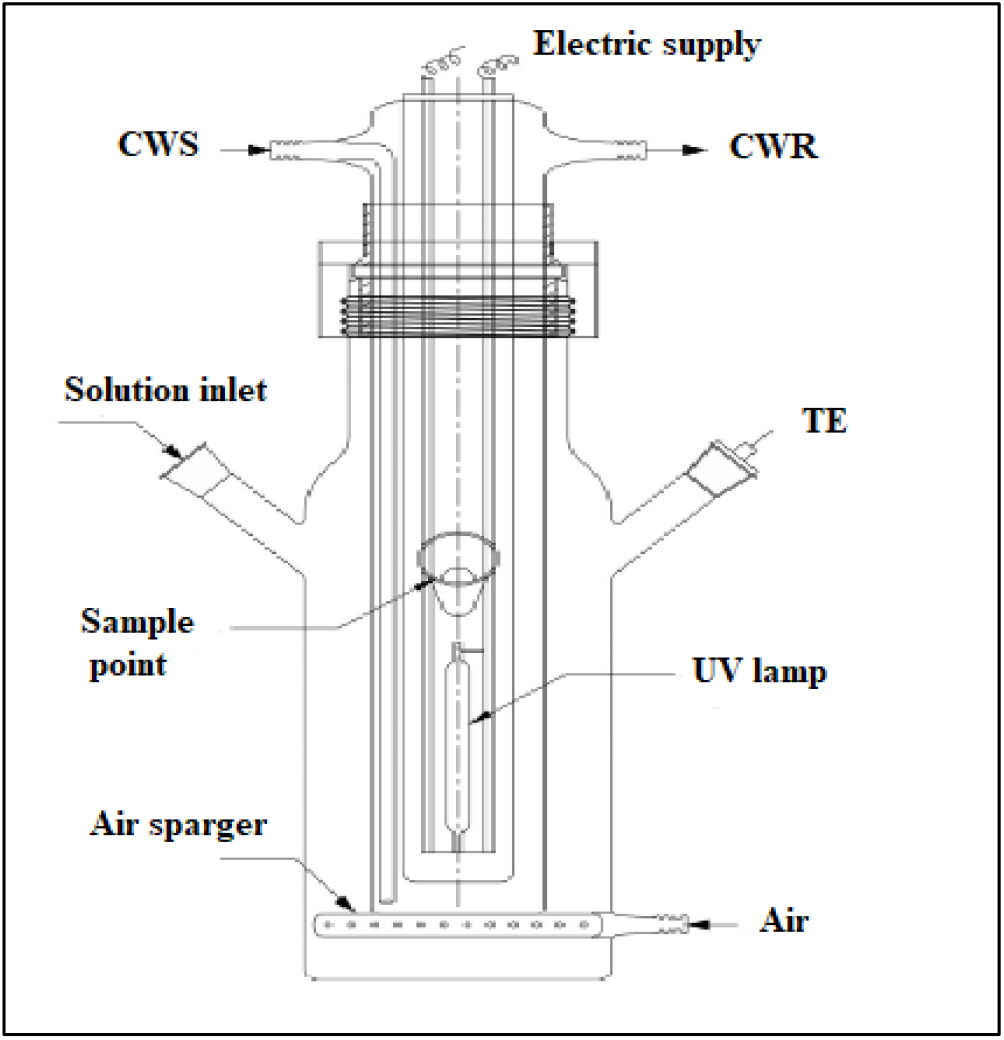
Setup for UV/ozonation and its hybrid processes.

### 2.2 Sampling of sewage water sample and sampling site

Raw sewage water samples were collected from the inlet of the Sewage Treatment Plant (Phytorid-STP) located at CSIR-National Chemical Laboratory (CSIR-NCL) campus, Pune, Maharashtra, India (18°32’31.2”N 73°48’43.2”E). The STP receives around 0.15 MLD of wastewater daily from Colony residents, hostel. Raw sewage water sample was collected following Standard Operating Procedure of wastewater surveillance by Center for disease control and prevention (CDC, USA) in a sterile container and stored at 4°C until it gets subjected to the following disinfection techniques. 20 litres of raw sewage water samples were collected for primary treatment and secondary treatment on 27^th^ July 2022 and 18^th^ September 2022, respectively. Whereas for evaluating the efficacy of the six best AOP techniques, three different sewage water samples (20 litres each) were collected on 30^th^ August 2022, 1^st^ September 2022, and 2^nd^ September 2022.

### 2.3 Treatment procedure

#### 2.3.1 Hydrodynamic cavitation and hybrid hydrodynamic cavitation technique

The known quantity of sewage water collected from STP was fed to the effluent holding tank (EHT), as shown in Fig. 2. The physiochemical characteristics like Total Organic Carbon (TOC), Total dissolved solids (TDS), Dissolve Oxygen (DO), and pH were estimated before and after AOPs treatment of raw sewage water. The sewage water was circulated from the EHT through the line consisting of a venturi device and back into the EHT using a centrifugal pump. The known quantity of ozone gas was introduced at the venturi throat depending on the requirement of disinfection techniques. In the case of hybrid techniques such as ozonation/H_2_O_2_/HC and ozonation/HC ozone gas is induced at the venturi throat, while in the case of HC and HC/H_2_O_2_ ozone gas is not induced. The sewage water was kept in recirculation mode for 90 min for treatment and the samples were taken and subsequently analysed for their physiochemical characteristics along with viral load. TOC also was measured using total organic carbon analyser TOC-L (Shimadzu model 00114). The pH, TDS, and DO were measured using electrodes provided by HANNA instruments (HI5521 and HI5522).

#### 2.3.2 Ozonation and hybrid techniques

Fig. 3 shows a schematic of the annular glass reactor used for UV, ozonation, and hybrid techniques such as UV/ H_2_O_2_, UV/ozonation, UV/ozonation/ H_2_O_2_, and ozonation/ H_2_O_2_. A known quantity of sewage water (500 ml) was added to the annular glass reactor having an annular space volume of 1 litre. The UV lamp of 80 W, having a wavelength of 254 nm, is placed in the inner quartz tube hollow candle, which will be used for UV and hybrid techniques such as UV/H_2_O_2_, UV/ozonation, UV/ozonation/H_2_O_2_. The reaction temperature was constantly maintained by circulating the chilled water from the annular space between the lamp with the help of JULABO chiller FP-50 MA. The ozone gas with the desired flow rate was supplied through a ring sparger situated at the bottom of the reactor. The sewage water samples were taken from the reactor to estimate physiochemical characteristics and viral load. The disinfection experiment was performed for 90 min.

### 2.4 Isolation of Total Nucleic Acids

Wizard® Enviro Wastewater TNA kit (Promega Corp., USA) was used to purify Total Nucleic Acid (TNA) from the raw sewage water (unpasteurised) as well as AOPs treated water samples at the same time to compare the difference in the viral concentration. TNA was isolated using the manufacturer protocol, which consists of 2 Steps: In the first step, TNA is captured on PureYield™ Midi Binding Column (Promega Corp., USA) and then eluted in 1 ml of pre-warmed (60°C) nuclease-free water (NFW); in the second one eluted TNA was further purified using a Mini spin column and concentrated in 40 µL volume.

### 2.5 Viral Reverse Transcriptase-quantitative PCR assay (RT-qPCR)

#### 2.5.1 Quantification of SARS-CoV-2 RNA

The isolated TNA was subjected to SARS-CoV-2 RNA screening using GenePath Dx CoViDx One v2.1.1TK-Quantitative multiplex RT-qPCR kit (Achira Labs, India). This kit targets nucleocapsid (N), RNA-dependent RNA Polymerase (RdRp), and Envelope (E) regions of the SARS-CoV-2 genome along with human control gene (RNAase P). 15 µL of amplification reaction was composed of 10 µL of reaction master mix and 5 µL TNA. NFW was used as a no-template control (NTC), and extraction control was analysed with each plate. Experiments were performed on 7500 Fast Real-Time PCR system (Applied Biosystems, Foster City, USA) with the following cycling conditions: Pre-incubation for 5 minutes at 37°C, Reverse transcription for 7 minutes at 52°C, R.T. inactivation for 3 minutes 30 seconds at 95°C, and 40 cycles of denaturation at 5 seconds at 95°C and extension 35 seconds at 35°C. RT-qPCR was performed in duplicates for each sample. The quantitation of the SARS-CoV-2 viral load in the samples was done using the Covid-19 Viral Load Calculation Tool (RUO).

#### 2.5.2 Quantification of PMMoV RNA

Quantification of PMMoV RNA was screened using the GenePath Dx Wastewater monitoring for Covid-19 (RUO) (Achira Labs, India) on a 7500 Fast Real-Time PCR system (Applied Biosystems, Foster City, USA). Three standards of PMMoV (of concentration 5000 copies/µL, 500 copies/µL, 50 copies/µL) were amplified along with experimental samples to construct a standard curve for PMMoV quantification. 5 µL of extracted TNA was mixed with 10 µL reaction master mix to make 15 µL amplification reaction. 5 µL of NFW was used as an NTC. Reactions were performed with the following cycling conditions: Pre-incubation for 5 minutes at 37°C, Reverse transcription for 7 minutes at 52°C, R.T. inactivation for 3 minutes 30 seconds at 95°C, and 40 cycles of denaturation at 5 seconds at 95°C and extension 35 seconds at 35°C. RT-qPCR was performed in duplicates for each sample. The concentration of PMMoV was estimated using the standard curve.

## 3. Results and Discussion

SARS-CoV-2 viral load in sewage water started to rise from the first week of July 2022. The escalated viral load in sewage water from the inlet of STP was needed to evaluate the best sewage water treatment techniques. Therefore, these experiments were accomplished from August 2022 to September 2022, when the viral load of sewage water was elevated. The disinfecting techniques like hydrodynamic cavitation (HC), UV and ozonation, as well as their hybrid combinations like HC/O_3_, HC/O_3_/H_2_O_2_, HC/H_2_O_2,_ O_3_/UV, UV/H_2_O_2_, UV/H_2_O_2_/O_3_ and O_3_/H_2_O_2_ for reducing the SARS-CoV-2 viral load from sewage water, were used in this study. The experiments were divided into three parts: 1) Primary screening of AOPs and hybrid AOPs, 2) Variation of ozone dose for optimum efficiency, 3) Treatment of three different sewage water samples using selected six best AOP techniques.

Around ten AOPs, including hybrids techniques were screened for effective disinfection of the SARS-CoV-2 from raw sewage water (Fig. 4). The ozone flow rate of 8 gm/hr during the initial screening was kept constant. TNA was isolated from a sewage water sample (raw and treated) and subjected to RT-qPCR, which were analysed by cycle threshold (Ct) and viral load. AOP treated sewage water having Ct value < 35 for any two target genes of SARS-CoV-2 were considered SARS-CoV-2 Positive (according to the kit’s instruction), and they were less efficient in reducing SARS-CoV-2 from sewage water (Supplementary Table: 1).

**Fig. 4.**
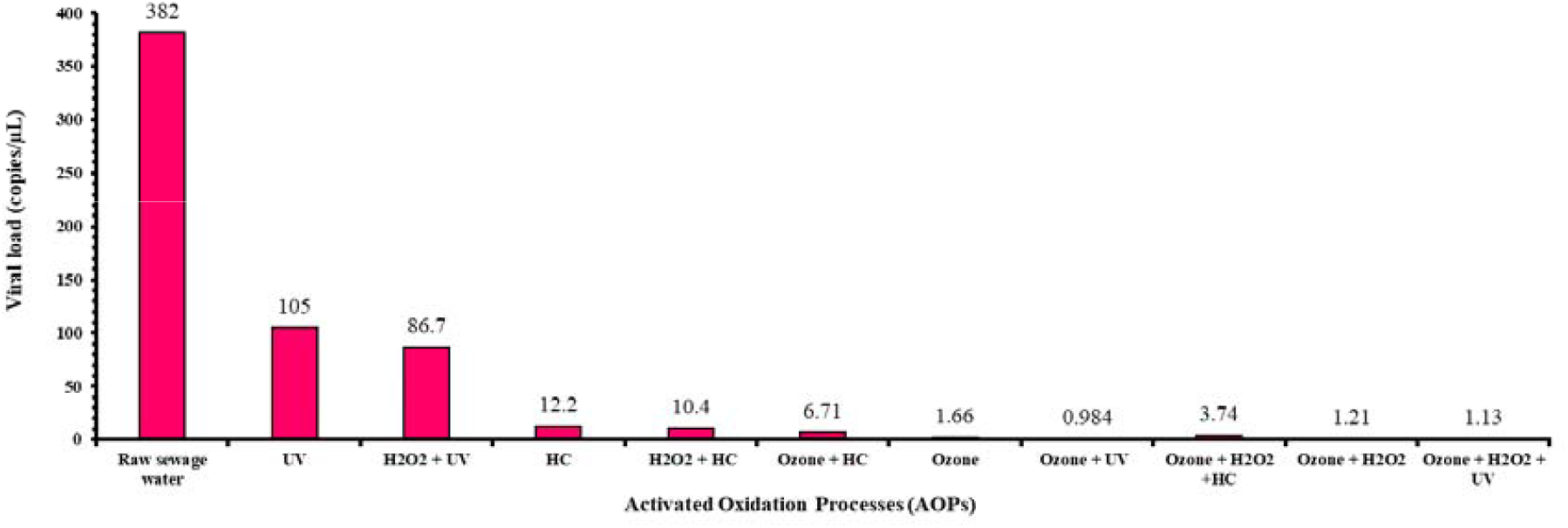
Bar plot illustrating evaluation of disinfecting efficiency of different AOPs for reduced viral load.

The viral load of raw sewage water used in the initial screening was 382 copies/µL. During the initial screening, UV was used as one of the disinfection techniques because it can eradicate biological pollutants from wastewater (Darnell et al., 2004; Parsa et al., 2021) and SARS-CoV-2 from infected surfaces like doors and handles of windows (Hadi et al., 2020; Kuzniewski, 2021). UV light of wavelength 254 nm was used in the treatment process as it is widely used for disinfection purposes (Singh et al., 2021). Therefore, eradication of SARS-CoV-2 RNA was done by exposing sewage water to UV for 90 minutes, giving a 75.5% reduction in RNA concentration which is 105 copies/µL. SARS-CoV-2 RNA was reduced to 86.7 copies/µL with disinfecting efficiency of 77.3% when UV was combined with hydrogen peroxide (400 mg/l); the synergetic effect of the combination of UV/H_2_O_2_ lead to more generation of hydroxyl radical which resulted in high disinfecting efficiency (Ibrahim et al., 2021). Dular et al., 2016 studied sewage water treatment using hydrodynamic cavitation technique and found that it reduces waterborne enteric viruses from sewage water treatment plants. Similarly, Bui et al., 2019 also investigated HC as an efficient and economical treatment technique that generates no by-products. In the present study, HC treatment of sewage water for 90 minutes scaled down the SARS-CoV-2 RNA viral load to 12.2 copies/µL with 96.80% disinfecting efficiency. But when HC was combined with hydrogen peroxide (400 mg/l) and Ozone (8 gm/hr) to treat the sewage water, the RNA concentration was reduced to 10.4 copies/µL and 6.71 copies/µL, respectively. Besides this, a hybrid of HC, ozone (8 gm/hr), and hydrogen peroxide (400 gm/l) brought the SARS-CoV-2 RNA concentration down to 3.7 copies/µL with 99.02% disinfecting efficiency because the combination of HC with other AOPs are more effective for sewage water treatment as the number of hydroxyl radical generated are more due to the addition of hydrogen peroxide and ozone. (Gogate et al., 2020; Thanekar and Gogate, 2019). To the best of our knowledge, this is the first study to implicate hydrodynamic cavitation for reducing SARS-CoV-2 viral load from sewage water. SARS-CoV-2 RNA concentration in sewage water was scaled down to 1.66 copies/µL when treated with ozone at an 8 gm/hr flow rate for 90 minutes. The combination of ozone (8 gm/hr) with UV and hydrogen peroxide (400 mg/l) was also used to treat sewage water, resulting in the diminution of SARS-CoV-2 RNA concentration to 0.984 copies/µL and 1.21 copies/µL, respectively. In addition, a hybrid treatment of ozone (8 gm/hr), UV and hydrogen peroxide (400 mg/l) was also used to treat sewage water and decline the SARS-CoV-2 RNA concentration to 1.13 copies/µL. All ozone and ozone based AOPs sewage water techniques shown more than 99.00% disinfecting capability, as it is highly reactive to viruses because of its oxidising property, and it can eradicate enveloped as well as non-enveloped viruses (Martins et al., 2021; Murray et al., 2008; Thanekar and Gogate, 2019; Young et al., 2020).

Variations of ozone dose were given to the six most effective techniques from the primary screening. Two variations were designed 8 gm ozone/hr and 15 gm ozone/hr for 90 minutes. In this screening, ozonation/UV, ozonation/UV/H_2_O_2,_ HC/ozonation, and ozonation/HC/H_2_O_2_ proved to be most effective when the ozone dose was 15 gm/hr (Fig. 5). The ozone flow rate of 8 gm/hr worked best for ozonation and ozonation/ H_2_O_2_. However, both ozone flow rate gives a desirable reduction in the SARS-CoV-2 RNA concentration (Fig. 5). These six best AOP techniques were further tried to treat three different sewage water samples collected on different days. The purpose of the treatment of three different sewage water samples was to certify the efficacy and consistency of all selected six best AOP techniques for the reduction of SARS-CoV-2 RNA concentration. It was found that all treatment techniques show SARS-CoV-2 RNA viral load reduction to the desired level (Fig. 6).

**Fig. 5.**
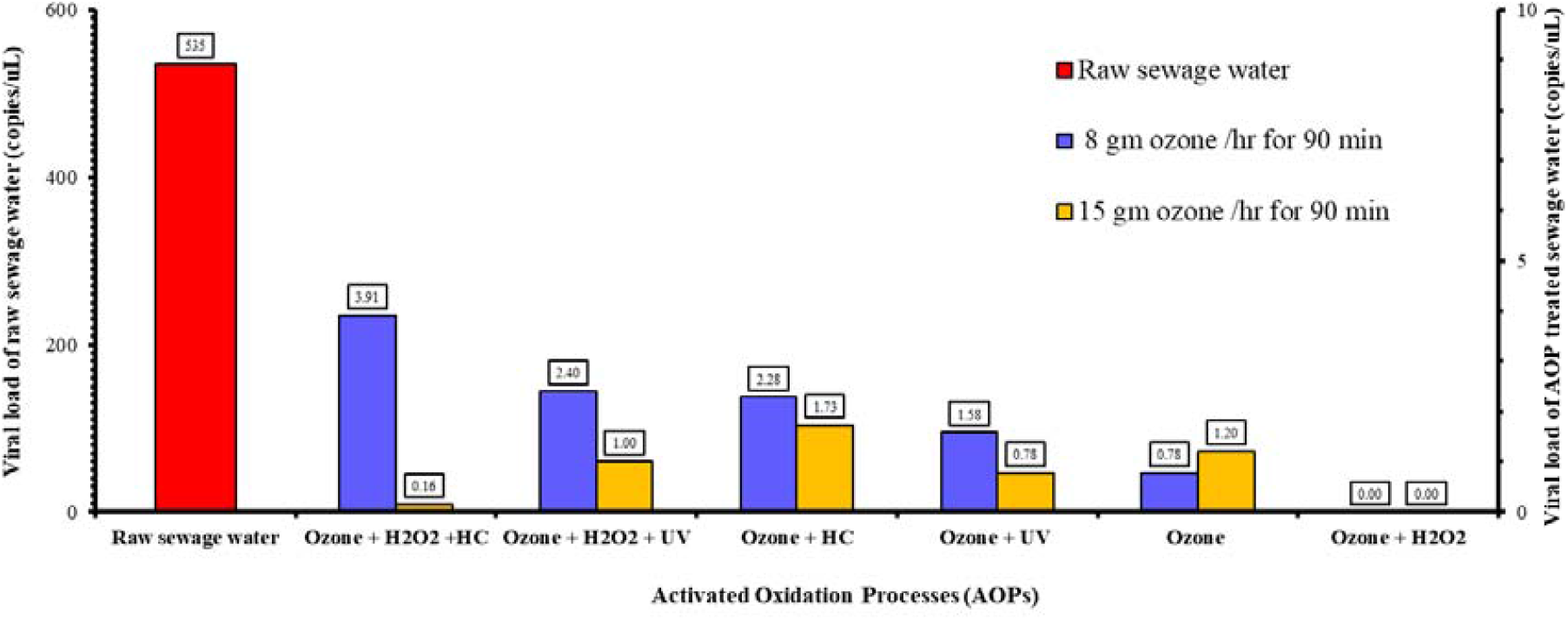
Variations of initially screened treatment techniques using raw sewage water samples. The Y-axis at the left shows the viral load (copies/µL) of raw sewage water (red bar), and the secondary Y-axis axis depicts the viral load (copies/µL) of AOP treated sewage water.

**Fig. 6.**
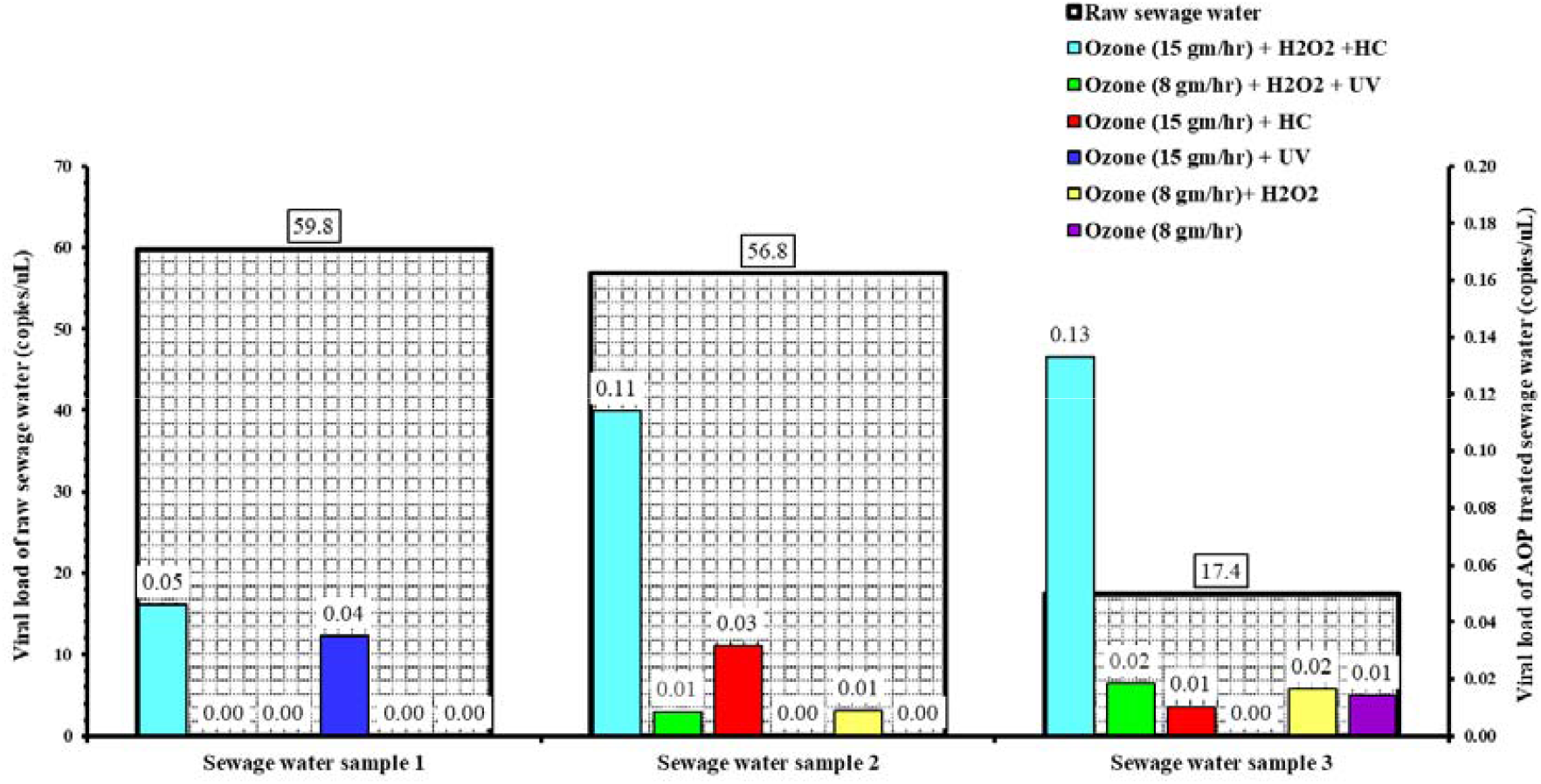
Graphical representation of the SARS-CoV-2 viral load reduction for the three different sewage water samples collected on different days using six best AOP techniques. The Y-axis at the left shows the viral load (copies/µL) of raw sewage water (checkered pattern bar), and the secondary axis represents the viral load (copies/µL) of AOP treated water.

Additionally, PMMoV RNA reduction was also evaluated for the three different sewage water samples collected on different days using the six best AOP techniques. PMMoV is the most abundant RNA virus in human faeces and an indicator of faecal contamination (Rosario et al., 2009). GenePathDx Wastewater monitoring for Covid-19 (RUO) (Achira Labs, India) was used to evaluate PMMoV RNA reduction and compared with raw sewage water sample (Fig. 7). PMMoV RNA was more than 1900 copies/µL in raw sewage water got reduced to <600 copies/µL after AOPs treatment, hence we can say that these treatment techniques are also effective in reducing the faecal contamination along with SARS-CoV-2 RNA from sewage water (Canh et al., 2022).

**Fig. 7.**
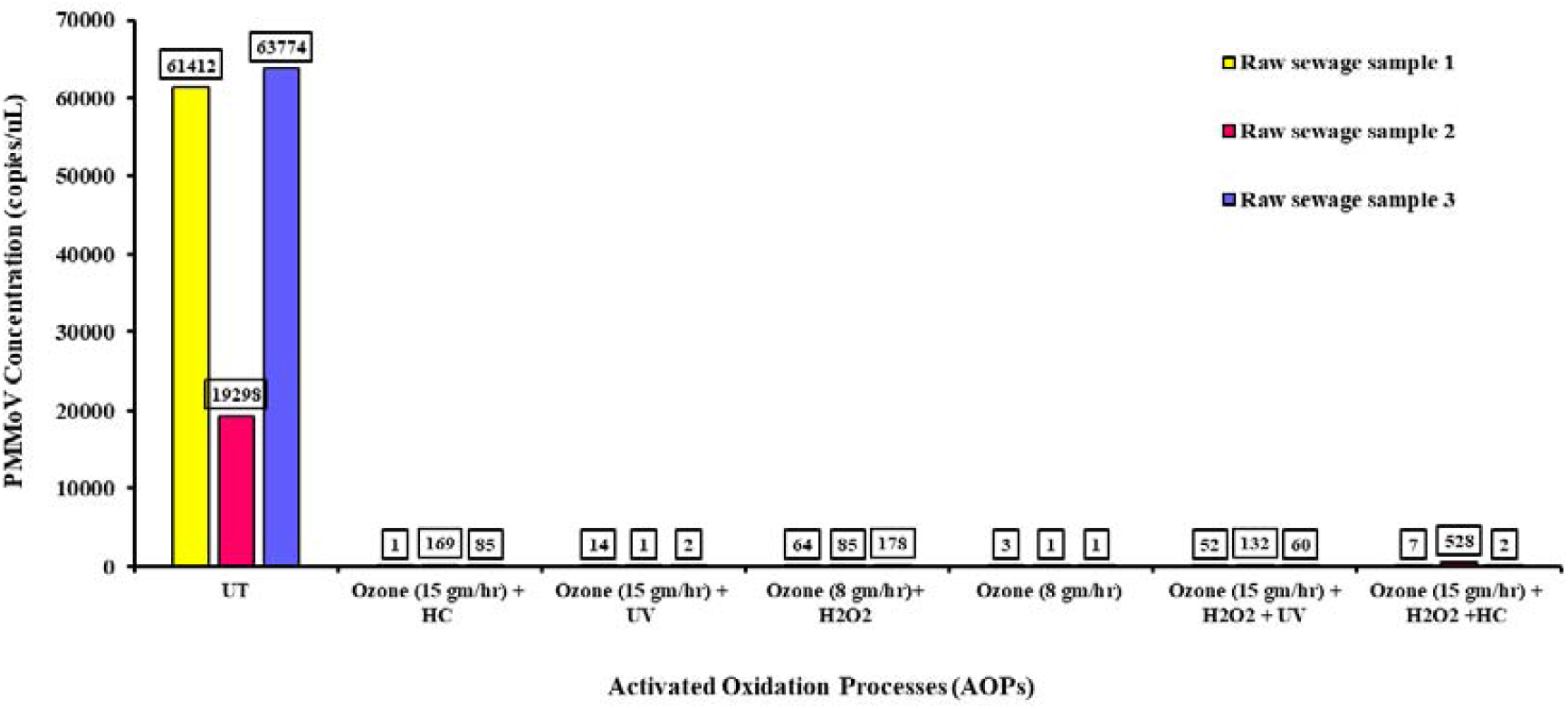
PMMoV RNA (copies/µL) reduction. After treatment of the three different sewage water samples collected using the six best AOP techniques.

The physicochemical characteristics of raw sewage water before and after the AOPs were estimated and shown in Table 2. The TOC of the treated sewage water decreases from 27 mg/L to below 5 mg/L for all treatment techniques (Table 2). This reflects a direct correlation between reduction in SARS-CoV-2 RNA, PMMoV, and other microbes in the sewage water and reduction in total organic carbon load as all these viruses and bacteria contain organic carbons (Li et al., 2022; Yu et al., 2022). Also, the dissolved oxygen (DO) level of treated sewage water has increased from 5 mg/L to 15 mg/L, which is a good indicator of water quality. Aquatic life depends on sufficient oxygen to live; hence, a drop in dissolved oxygen below the limit will result in the loss of fish and plants. Therefore, water quality can be estimated by measuring dissolved oxygen level. As per the Environmental Protection Agency (EPA) guideline, if dissolved oxygen concentration reaches 3 mg/L, it is considered to be danger zone for aquatic life and levels below 1 mg/L are unfavourable to aquatic life, whereas dissolved oxygen level concentration 8-9 mg/L support all aquatic life (fish and plants) (“Indicators: Dissolved Oxygen | US EPA,” 2022; Sofia, 2020). The above AOPs were performed for the first time to reduce SARS-CoV-2 from sewage water, and further studies are essential to implement these techniques for the treatment of sewage water at actual STP.

**Table 2.** Physical properties of untreated and treated sewage water using AOPs.

| Physiochemical<br>Parameters | TOC<br>(ppm) | TDS (ppm) | DO (ppm) | pH |
| --- | --- | --- | --- | --- |
| Raw sewage water | 27.06 | 336.4 | 5.9 | 7.5 |
| HC/Ozonation | 1.13 | 253.3 | 8.38 | 9 |
| UV/Ozonation | 5.58 | 261.3 | 12.7 | 8.7 |
| H<sub>2</sub>O<sub>2</sub>/Ozonation | 1.17 | 238.7 | 13.77 | 8.8 |
| Ozonation | 1.39 | 212.3 | 13.8 | 8.8 |
| UV/Ozonation / H<sub>2</sub>O<sub>2</sub> | 3.12 | 238.4 | 14.62 | 9 |
| HC/Ozonation/ H<sub>2</sub>O<sub>2</sub> | 3.37 | 268.8 | 9.37 | 8.4 |

## Conclusions

This is the first study showing the effective use of AOPs, including hydrodynamic cavitation (HC), ozonation, and hybrid AOPs shows more than 95% SARS-CoV-2 virus reduction. It was also found that ozone and ozone based hybrid AOPs techniques showed 99% effectiveness in disinfecting SARS-CoV-2. These AOP techniques were also found to effectively reduce PMMoV RNA concentration, a faecal indicator from sewage water. Although this is a preliminary study with a limited number of samples, the observations and experimental evidence highlighted the importance of AOP techniques in reducing SARS-CoV-2 viral load in sewage water. The results of our investigation could be instrumental for further studies dealing with the prospection of AOPs for the reduction of other viruses and pathogens from the sewage water.

## Data Availability

All data produced in the present work are contained in the manuscript

## Acknowledgements

Authors are thankful to the Director, CSIR National Chemical Laboratory for support. This work funded by CSIR through project E3OW (MLP102326B) and National Chemical Laboratory, Pune (MLP 038526). Authors would like to express thanks to Engineering Division of National Chemical Laboratory for support during sample collection. Manuscript has been checked for plagiarism using iThenticate licensed version.

