## Supplementary Information for "Comprative evaluation of Advanced Oxidation Processes (AOPs) for reducing SARS-CoV-2 viral load from campus sewage water"

**Supplementary Table 1:** Primary Screening of AOPs (ND= Not detected, Ct < 35 positive, Ct> 35 negative for SARS-CoV-2)

| Treatment Method | Gene Targets | | | |
| --- | --- | --- | --- | --- |
|  | RdRP Gene | N Gene | E Gene | RNaseP |
| Raw sewage water | 28.49 | 26.98 | 29.3 | 34.15 |
| UV+ Ozone | 39.68 | 36.78 | ND | ND |
| H_2_O_2_+Ozone | 39.99 | ND | ND | 36.85 |
| Ozone | 39.2 | 35.25 | ND | 36.32 |
| UV + H_2_O_2_ | 31.68 | 29.11 | 32.46 | 35.96 |
| UV+ Ozone + H_2_O_2_ | 40.01 | 37.41 | 37.62 | 36.45 |
| UV | 31.16 | 29.38 | 31.87 | 36.46 |
| Cavitation + Ozone + H_2_O_2_ | 37.5 | 33.6 | 37.41 | ND |
| Cavitation + Ozone | 35.26 | 31.35 | 35.08 | 32.56 |
| Cavitation | 35.4 | 32.13 | 36.4 | 36.86 |
| Cavitation + H_2_O_2_ | 34.88 | 32.75 | 35.43 | 35.91 |

**Supplementary Table 2.** Secondary Screening of AOPs with variation of ozone dose (ND= Not detected, Ct < 35 positives, Ct> 35 negatives for SARS-CoV-2)

| Treatment | Ozone dose (gm/hr) | Gene Targets | | | |
| --- | --- | --- | --- | --- | --- |
|  |  | RdRP Gene | N Gene | E Gene | RNaseP |
| Raw sewage water | NA | 28.33 | 27.28 | 32.29 | 31.73 |
| HC/Ozonation/H_2_O_2_ | 8 | 35.12 | 30.66 | 37.37 | 32.08 |
| HC/Ozonation/H_2_O_2_ | 15 | ND | 35.56 | ND | ND |
| HC/Ozonation | 8 | 37.6 | ND | 40.66 | 37.84 |
| HC/Ozonation | 15 | 38.96 | ND | ND | ND |
| UV/Ozonation | 8 | 39.93 | 38.75 | 37.63 | 36.31 |
| UV/Ozonation | 15 | 39.23 | 39.5 | ND | ND |
| H_2_O_2/_Ozonation | 8 | ND | ND | ND | ND |
| H_2_O_2/_Ozonation | 15 | ND | ND | ND | ND |
| Ozonation | 8 | 40 | ND | 40.38 | ND |
| Ozonation | 15 | 39.55 | ND | ND | ND |
| UV/Ozonation/H_2_O_2_ | 8 | 38.01 | 36.41 | 37.62 | 36.45 |
| UV/Ozonation/H_2_O_2_ | 15 | 39.76 | 35.82 | 39.73 | 37.1 |

**Supplementary Table 3.** Three different sewage water samples were treated using six best AOPs (ND= Not detected, Ct < 35 positives, Ct> 35 negatives for SARS-CoV-2)

| Treatment | Ozone dose | Sewage water sample 1 | | | | Sewage water sample 2 | | | | Sewage water sample 3 | | | |
| --- | --- | --- | --- | --- | --- | --- | --- | --- | --- | --- | --- | --- | --- |
|  |  | RdRP Gene | N Gene | E Gene | RNaseP | RdRP Gene | N Gene | E Gene | RNaseP | RdRP Gene | N Gene | E Gene | RNaseP |
| Raw sewage water | NA | 31.32 | 29.12 | 34.9 | 30.17 | 31.3 | 29.23 | 35.94 | 32.42 | 33.83 | 31.71 | 34.51 | 31.53 |
| HC/Ozonation | 15 | 38.71 | ND | ND | 38.71 | ND | 39.26 | ND | 35.15 | ND | 40.97 | ND | 36.54 |
| UV/Ozonation | 15 | ND | 39.1 | ND | ND | ND | ND | ND | 38.4 | ND | ND | ND | 39.27 |
| H_2_O_2/_Ozonation | 8 | ND | ND | ND | 38.41 | ND | 41.19 | ND | 35.95 | ND | 40.24 | ND | 32.81 |
| Ozonation | 8 | ND | ND | ND | ND | ND | ND | ND | ND | ND | 40.46 | ND | ND |
| UV/Ozonation/H_2_O_2_ | 15 | ND | ND | ND | 36.35 | ND | 41.24 | ND | 34.28 | ND | 40.08 | ND | 34.55 |
| HC/Ozonation/H_2_O_2_ | 15 | ND | 38.69 | ND | 37.86 | 41.56 | ND | ND | 35.12 | ND | 37.09 | ND | 37.06 |
